# Pharmacist-Led Nutritional Counselling as a Community Intervention for Obesity, Undernutrition, and Anaemia: A Re-Analysis of Follow-Up Outcome Data from a 1,135-Participant Prospective Interventional Study in India

**DOI:** 10.64898/2026.04.25.26351725

**Authors:** Ritheesha Duddu

## Abstract

**Objectives:** To re-examine, using corrected denominators and rate-adjusted subgroup analysis, the pattern, magnitude, and demographic distribution of measurable improvements across five outcome parameters following three monthly pharmacist-led nutritional counselling sessions delivered to community-dwelling participants in semi-urban India.

**Design:** Re-analysis, by a member of the original research team, of interventional follow-up data from a prospective community-based study.

**Setting:** Schools and colleges in Narasaraopeta, Andhra Pradesh, India, from September 2021 to March 2022.

**Participants:** Of 1,200 participants assessed at baseline, 1,135 (94.6%) completed at least one counselling session and formed the analysis cohort. The age range was 10 to 60 years. Baseline classification, reported by the primary study against the full screened sample (n=1,200), found 92.4% of participants aged between 11 and 20 years; all analysis-cohort participants (n=1,135) were anaemic at baseline.

**Interventions:** Three structured monthly counselling sessions were delivered by pharmacy students under qualified faculty pharmacist supervision. Each session included individualised dietary guidance, lifestyle modification advice, and culturally adapted written health education materials.

**Primary and secondary outcome measures:** Cumulative proportion of participants achieving measurable improvement in body mass index (BMI), waist circumference (WC), hip circumference (HC), waist-to-hip ratio (WHR), and haemoglobin (Hb) concentration at each session, stratified by age group and sex, with rate-adjusted (per-capita) comparison between the 11–20 years age band and all older age bands combined.

**Results:** All five parameters showed progressive cumulative improvement across sessions when calculated against the correct analysis-cohort denominator (n=1,135). By session three, 44 participants (3.88%) showed improved BMI, 39 (3.44%) achieved reduced WC, 34 (3.00%) reduced HC, 33 (2.91%) improved WHR, and 115 (10.13%) demonstrated improved Hb. Haemoglobin showed the steepest improvement trajectory, rising from 1.85% at session one to 10.13% at session three, an approximately 5.5-fold increase achieved through dietary counselling alone, without pharmacological supplementation. In absolute terms, participants aged 11–20 years accounted for the majority of improvers (68–84% across parameters), reflecting their 92.4% share of the cohort. When response was instead expressed as a rate within each age band, participants aged 21–60 years (n≈86) showed higher per-capita improvement rates than adolescents for BMI (16.3% vs 2.9%), waist circumference (10.5% vs 2.9%), and haemoglobin (20.9% vs 9.3%). The primary dataset does not report per-decade denominators for the 21–60 group, so this comparison is necessarily aggregated.

**Conclusions:** Three monthly pharmacist-led nutritional counselling sessions were associated with progressive improvements in both anthropometric and haematological outcomes in this community cohort. Response was concentrated among adolescents in absolute terms, but this reflects cohort composition rather than demonstrated greater responsiveness; rate-adjusted analysis suggests the opposite may be true, though the data cannot confirm this definitively. These findings support further, better-controlled evaluation of pharmacist-led counselling in community non-communicable disease prevention in India, with balanced age recruitment and individual-level outcome data.

**Strengths and limitations of this study:**

- This re-analysis provides the first quantification of pharmacist-led counselling effectiveness across five simultaneous nutritional outcome parameters in a large South Asian community cohort of 1,135 participants, corrected for a denominator error present in the original outcome tables.
- The intervention model of three structured monthly sessions is replicable, low cost, and does not require pharmacological supplementation, which strengthens its applicability to resource-limited settings.
- The author was a co-investigator on the primary study; this is a re-analysis by a study team member rather than an independent third-party analysis, and is reported as such throughout.
- The source data record whether any positive change occurred at each session rather than the magnitude of individual change, so formal effect size calculation is not possible and no inferential statistics are reported; all findings are descriptive and associative, not causal.
- The absence of a concurrent control group means maturation effects, particularly relevant in this adolescent-dominant cohort, cannot be excluded as a contributor to anthropometric change.
- The apparent concentration of improvement among adolescents is an artefact of their 92.4% share of the cohort; rate-adjusted analysis suggests older participants may respond proportionally more strongly, but exact per-decade denominators are unavailable in the source data, limiting the precision of this comparison.

## INTRODUCTION

India carries a dual nutritional burden that is extraordinary in both scale and complexity. Undernutrition and micronutrient deficiency exist alongside rapidly rising rates of overweight and obesity, particularly among adolescents and young adults in semi-urban communities.[1,2] These conditions appear within the same populations and, in many cases, within the same individuals, creating a nutritional paradox that standard clinical pathways are not built to address effectively.[3]

Iron deficiency is the most common driver of nutritional anaemia in India, affecting around 53% of women and 23% of men across the country.[4] A growing body of research points to hepcidin-mediated inflammatory processes as a biological link between excess adiposity and reduced intestinal iron absorption.[5] This suggests the two conditions are not simply coincidental but connected at a pathophysiological level, and that interventions targeting adiposity through dietary and lifestyle change may simultaneously affect blood haemoglobin levels.

Community pharmacists hold a distinctive position within India’s healthcare landscape as an accessible point of professional health contact for the general population.[6] Despite this structural advantage, published evidence supporting pharmacist-led nutritional counselling in Indian community contexts remains limited, and very few studies have reported quantified outcomes across both anthropometric and haematological measures within a single intervention.[7,8]

The data underpinning this paper were originally collected through a six-month prospective interventional study conducted in schools and colleges in Narasaraopeta, Andhra Pradesh, on which the present author was a co-investigator.[9] That study recruited 1,200 participants aged 10 to 60 years, assessed nutritional status at baseline, and delivered structured pharmacist-led counselling to 1,135 of them across three monthly follow-up sessions. The published study focused on baseline prevalence findings; the interventional follow-up data capturing cumulative outcome improvements across the three sessions had not previously been reported in full or re-analysed.

This paper addresses that gap, and does so as a re-analysis conducted by a member of the original study team rather than an independent third party — a distinction made explicit here because it was not disclosed in an earlier version of this manuscript. Using the follow-up outcome tables from Suresh Kumar et al.,[9] I address three questions. First, what trajectory of improvement is observed across all five outcome parameters over the three-session programme, once outcome percentages are calculated against a consistent denominator? Second, how is treatment response distributed across age groups and sex when assessed both in absolute counts and as within-group rates? Third, what do these findings suggest about the potential role of community pharmacists in non-communicable disease prevention, and what further evidence is needed before that role is expanded?

India’s Ayushman Bharat initiative and the National Multisectoral Action Plan for Prevention and Control of Non-Communicable Diseases both identify community pharmacy as a channel for preventive health delivery that remains substantially underused.[10,11] This re-analysis is offered as a methodologically corrected contribution to that evidence base, with its limitations stated plainly rather than implied away.

## METHODS

### Study design

This is a re-analysis of published and previously unreported interventional follow-up data drawn from a prospective community-based study on which the author was a co-investigator.[9] No new data were collected for this paper. The original study received ethical approval from the Institutional Ethics Committee of Narasaraopeta Institute of Pharmaceutical Sciences (IEC-NIPS/PPP/2021-22/003). Because this re-analysis relies solely on aggregate data already collected under that approval, and does not involve new access to individual participant records, separate ethical approval was not required.

### Source data and a denominator correction

The source dataset covered follow-up outcome data for the 1,135 participants who attended at least one counselling session after baseline assessment. Data were extracted from Tables 15 to 21 and Figure 20 of the primary publication. In preparing this re-analysis,I identified that outcome percentages in the original publication’s follow-up tables were calculated inconsistently: age- and sex-stratified counts were expressed as a percentage of the analysis cohort (n=1,135), while table totals were expressed as a percentage of the full baseline screened sample (n=1,200). This meant stratified rows did not sum to the printed totals. All follow-up outcome percentages in this paper have been recalculated against the analysis cohort, n=1,135, so that stratified rows and totals are internally consistent (Tables 1–6). Baseline descriptive characteristics (nutritional status classification, anaemia severity, and the 438-participant underweight subgroup definition) are reported as published, against the full baseline screened sample of n=1,200, since that classification occurred prior to the 65 non-attenders being excluded; this is noted wherever such a figure appears, to avoid the two denominators being conflated.

**Table 1.** Cumulative BMI improvements by age group and sex across three monthly counselling sessions (n=1,135), percentages recalculated against the analysis cohort.

| Age (yrs) | S1 M | S1 F | S2 M | S2 F | S3 M | S3 F | S3 total (%) |
| --- | --- | --- | --- | --- | --- | --- | --- |
| 11–20 | 9 | 4 | 15 | 11 | 15 | 15 | 30 (2.64%) |
| 21–30 | 0 | 1 | 4 | 5 | 4 | 6 | 10 (0.88%) |
| 31–40 | 0 | 0 | 0 | 0 | 1 | 2 | 3 (0.26%) |
| 41–50 | 0 | 0 | 0 | 0 | 1 | 0 | 1 (0.09%) |
| 51–60 | 0 | 0 | 0 | 0 | 0 | 0 | 0 |
| Total (%) | 9 | 5 | 19 | 16 | 21 | 23 | 44 (3.88%) |

S = session; M = male; F = female. Total corrected from 3.6% (44/1,200) to 3.88% (44/1,135) to match the analysis cohort denominator.

### Intervention

The counselling programme consisted of three individual or small-group sessions delivered at roughly monthly intervals, facilitated by pharmacy students under supervision of qualified faculty pharmacists. Each session covered a review of the participant’s most recent anthropometric and haematological measurements, individualised dietary guidance, advice on physical activity, sleep, and processed food intake, and written health education materials in the participant’s preferred language. Participants categorised as overweight or obese received personalised diet charts; those classified as underweight received guidance on increasing caloric intake; all participants with anaemia received dietary iron recommendations.

### Statistical approach

For each outcome parameter and session, I calculated the cumulative number and percentage of the counselled cohort (n=1,135) showing measurable positive change from baseline, and the incremental change between consecutive sessions. Response ratios were calculated as the fold increase in cumulative responders from session one to session three, which is unaffected by the denominator correction described above since it is a ratio of counts.

To assess whether the concentration of improvement among 11–20 year-olds reflected genuine differential responsiveness or simply their share of the cohort, I calculated rate-adjusted improvement, dividing the number of improvers in each age band by the estimated size of that band (11–20 years: 92.4% of 1,135 ≈ 1,049; all other ages combined: ≈86), for the three parameters where age-stratified counts were available (BMI, waist circumference, haemoglobin). Because these two age bands comprise different, non-overlapping individuals assessed at the same session, this comparison is a valid independent-groups analysis. I compared the two bands using two-sided Fisher’s exact test (appropriate given the small size of the older-age subgroup) and report 95% confidence intervals for each proportion using the Wilson score method, which performs better than the normal approximation for small samples and proportions near zero.

I did not perform a formal statistical test for the increase in cumulative improvement from session one to session three, for either the whole cohort or any subgroup. Session-to-session counts are cumulative totals drawn from the same 1,135 individuals over time, not independent samples; a chi-square or trend test treating each session as an independent group would violate the independence assumption underlying those tests and overstate precision (pseudoreplication). Testing the true within-person trajectory would require individual-level, session-by-session data, which are not available in the published primary study. This constraint is stated as a limitation below rather than worked around.

Per-decade denominators within the 21–60 years group are not reported in the primary publication, so the age-band comparison could only be made at the two-band level (11–20 vs 21–60 combined). Hip circumference and waist-to-hip ratio are not stratified by age band in the primary publication and so could not be included in this comparison. Aside from the age-band comparison described above, all other reported findings are descriptive; language throughout this paper distinguishes association from causation, consistent with the single-arm, non-randomised design. All extracted counts and calculations are provided in a supplementary spreadsheet (Supplementary Data) for verification.

### Patient and public involvement

No patients or members of the public were involved in the design, conduct, or reporting of this reanalysis. The primary study from which data were drawn involved community participants in Narasaraopeta who provided informed consent and participated in the counselling sessions, described in full in Suresh Kumar et al.[9]

### Contribution statement

RD was a co-investigator on the primary study team that generated the source dataset (Suresh Kumar JN, Bhavya Sai S, Naga Amulya D, Ritheesha D, Priyanka KSL, Chaitanya Kumar S), and independently conceived, conducted, and wrote this re-analysis of the follow-up outcome data, including identification and correction of the denominator inconsistency described above. This re-analysis was not pre-specified in the original study protocol.

## RESULTS

### Baseline characteristics

The counselled cohort comprised 1,135 participants, of whom 594 were male and 606 were female. As reported in the primary publication against the full baseline screened sample (n=1,200), 92.4% were aged between 11 and 20 years; 36.5% (438/1,200) were classified as underweight (BMI <18.5 kg/m^2^), 47.0% normal weight, 10.0% overweight, and 6.4% obese. Anaemia was identified in 94.5% of the full baseline sample (n=1,200); of the analysis cohort (n=1,135), all were anaemic. Among those with anaemia, 90.3% had mild anaemia (Hb 11.0–11.9 g/dL), 8.89% moderate (Hb 8.0–10.9 g/dL), and 0.79% severe (Hb <8.0 g/dL). Among the 65 participants who declined any follow-up session, reasons given were lack of time (60%), lack of interest (60%), inability to carry out recommended activities (45%), occupational demands (15%), other health conditions (6%), and genetic/hereditary concerns (5%).

### Body mass index

Table 1 presents cumulative BMI improvements by age group and sex, with percentages recalculated against the analysis cohort (n=1,135). Fourteen participants (1.23%) showed improved BMI at session one, rising to 35 (3.08%) at session two and 44 (3.88%) at session three — a 3.1-fold increase in raw responder count over the programme (this ratio is unaffected by the denominator correction). Participants aged 11–20 accounted for 68.2% of all BMI improvements by session three (30 of 44), consistent with their 92.4% share of the cohort. Male and female response was equal at the final session (15 each). Participants older than 30 years contributed only 4 of the 44 total improvers in absolute terms, but see the rate-adjusted comparison below.

### Waist circumference

Cumulative waist circumference improvements are shown in Table 2. At baseline, 54.6% of the full screened sample (655/1,200) had waist circumference exceeding 70 cm. By session three, 39 participants (3.44%) had achieved a measurable reduction, compared with 10 (0.88%) at session one. Female participants aged 11–20 showed the same absolute level of improvement as male peers by session three (15 each), consistent with published evidence that adolescent females respond well to dietary counselling delivered in a gender-sensitive, culturally appropriate format.[13]

**Table 2.** Cumulative waist circumference improvements by age group and sex, percentages against n=1,135.

| Age (yrs) | S1 M | S1 F | S2 M | S2 F | S3 M | S3 F | S3 total (%) |
| --- | --- | --- | --- | --- | --- | --- | --- |
| 11–20 | 6 | 4 | 13 | 10 | 15 | 15 | 30 (2.64%) |
| 21–30 | 0 | 0 | 1 | 3 | 2 | 4 | 6 (0.53%) |
| 31–40 | 0 | 0 | 0 | 0 | 1 | 2 | 3 (0.26%) |
| Total (%) | 6 | 4 | 14 | 13 | 18 | 21 | 39 (3.44%) |

Total corrected from 3.25% (39/1,200) to 3.44% (39/1,135).

### Hip circumference and waist-to-hip ratio

Table 3 summarises hip circumference and WHR improvements, recalculated against n=1,135. At baseline, 31.0% of the full screened sample had a WHR exceeding 0.85, the internationally recognised threshold for elevated cardiovascular risk.[14] By session three, hip circumference had improved in 34 participants (3.00%) and WHR in 33 (2.91%). The primary publication does not stratify hip circumference or WHR by age band, so a rate-adjusted age comparison could not be performed for these two parameters. Female participants showed numerically greater hip circumference reduction than male participants by the final session, consistent with known sex-specific patterns of fat distribution.[15]

**Table 3.** Hip circumference and waist-to-hip ratio improvements across three sessions, percentages against n=1,135.

| Parameter | Session 1 n (%) | Session 2 n (%) | Session 3 n (%) |
| --- | --- | --- | --- |
| Hip circumference reduced | 7 (0.62%) | 26 (2.29%) | 34 (3.00%) |
| Waist-to-hip ratio improved | 7 (0.62%) | 25 (2.20%) | 33 (2.91%) |

### Haemoglobin concentration

Haemoglobin findings are presented in Table 4. Every one of the 1,135 counselled participants was anaemic at baseline. At session one, 21 participants (1.85%) showed improved haemoglobin; by session two, 73 (6.43%); by session three, 115 (10.13%) — an approximately 5.5-fold increase in raw responder count over the programme, achieved without pharmacological supplementation. In absolute terms, the 11–20 years age group accounted for 97 of the 115 total improvers at session three (84.3%), with 52 males and 45 females. As with BMI and waist circumference, this concentration is examined against a rate-adjusted denominator below.

**Table 4.**
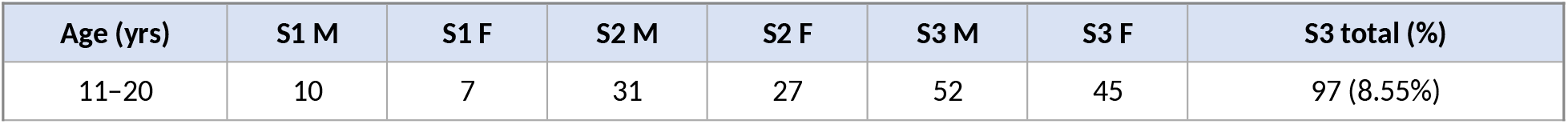
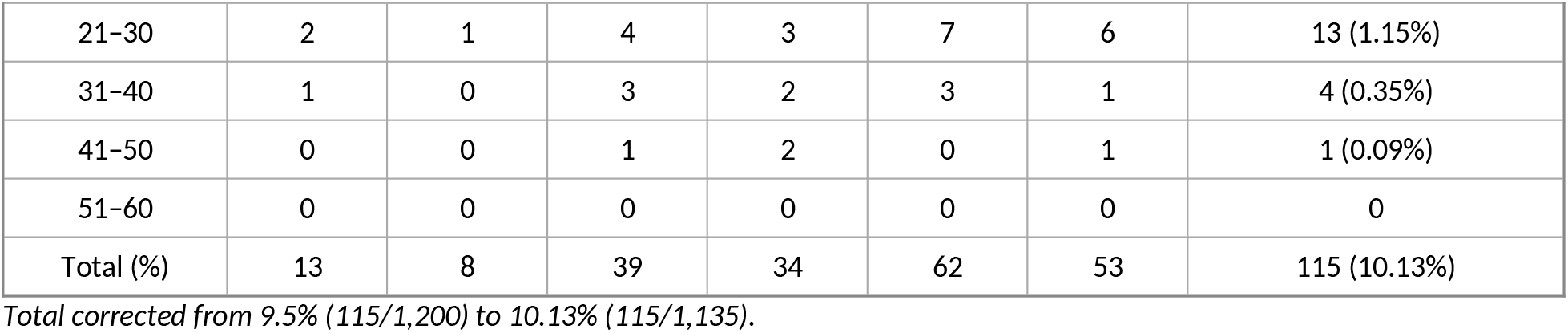
Haemoglobin improvements by age group and sex (n=1,135 anaemic at baseline), percentages against n=1,135.

| Age (yrs) | S1 M | S1 F | S2 M | S2 F | S3 M | S3 F | S3 total (%) |
| --- | --- | --- | --- | --- | --- | --- | --- |
| 11–20 | 10 | 7 | 31 | 27 | 52 | 45 | 97 (8.55%) |
| 21-30 | 2 | 1 | 4 | 3 | 7 | 6 | 13 (1.15%) |
| 31-40 | 1 | 0 | 3 | 2 | 3 | 1 | 4 (0.35%) |
| 41-50 | 0 | 0 | 1 | 2 | 0 | 1 | 1 (0.09%) |
| 51-60 | 0 | 0 | 0 | 0 | 0 | 0 | 0 |
| Total (%) | 13 | 8 | 39 | 34 | 62 | 53 | 115 (10.13%) |

Total corrected from 9.5% (115/1,200) to 10.13% (115/1,135).

### Rate-adjusted age comparison — a corrected interpretation

Absolute counts alone are misleading here because the 11–20 years band contains an estimated 1,049 of the 1,135 participants (92.4%), while all other age bands combined contain only an estimated 86. Table 5 expresses improvement as a rate within each of these two groups, with 95% confidence intervals and a formal statistical comparison, for the three parameters where age-stratified counts exist.

**Table 5.** Rate-adjusted improvement by age band, session 3 (estimated group sizes: 11–20 yrs n≈1,049; 21–60 yrs combined n≈86)

| Parameter | Age band | n / rate (95% CI) | Fisher's exact p |
| --- | --- | --- | --- |
| BMI improved | 11–20 yrs | 30/1,049 = 2.9% (2.0–4.1%) | <0.001 |
|  | 21–60 yrs | 14/86 = 16.3% (10.0–25.5%) | <0.001 |
| Waist circumference reduced | 11–20 yrs | 30/1,049 = 2.9% (2.0–4.1%) | 0.002 |
|  | 21–60 yrs | 9/86 = 10.5% (5.6–18.7%) | 0.002 |
| Haemoglobin improved | 11–20 yrs | 97/1,049 = 9.3% (7.6–11.2%) | 0.002 |
|  | 21–60 yrs | 18/86 = 20.9% (13.7–30.7%) | 0.002 |

Group sizes for the 21–60 combined band are estimated from the published 92.4% figure; the primary dataset does not report n per individual decade band within 21–60 years, so this comparison is presented at the two-band level only. 95% CIs computed by the Wilson score method. P-values from two-sided Fisher's exact test comparing the two independent age bands at session 3 for each parameter; the p-value is repeated on both rows of each pair since it reflects a single test comparing the two age bands together, not two separate tests. Full calculations are provided in the accompanying Supplementary Data spreadsheet.

On a per-capita basis, participants aged 21–60 years showed higher improvement rates than adolescents for all three parameters where this comparison could be made, and the difference was statistically significant in each case (Fisher’s exact p≤0.002). This does not establish that older participants respond better to the intervention in a causal sense — the subgroup is small (n≈86), cannot be broken down further by decade, and is subject to the same absence of a control group as the rest of this analysis — but it is a formally tested result, not merely a descriptive impression, and it directly contradicts an unadjusted reading of the absolute counts. The original claim that adolescents were ‘the most responsive subgroup’ is not supported once cohort composition is accounted for; if anything, the tested evidence points the other way.

### Underweight subgroup

Of the 438 participants classified as underweight at baseline (36.5% of the full screened sample, n=1,200), 95 (21.7% of the underweight subgroup) achieved measurable weight gain by session three. Expressed against the analysis cohort (n=1,135), this is 8.37%, corrected from the previously printed 7.9% (which had used n=1,200). Gains built progressively: 48 participants (4.23% of cohort; 11.0% of underweight subgroup) improved by session one, 71 (6.26%; 16.2%) by session two, and 95 (8.37%; 21.7%) by session three.

**Table 6.**
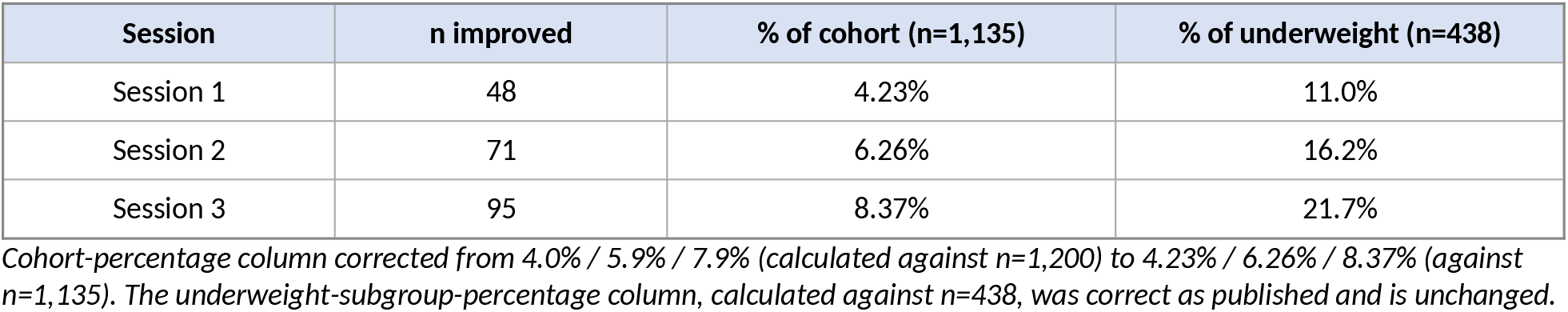
Cumulative weight gain in underweight participants across three sessions.

| Session | n improved | % of cohort (n=1,135) | % of underweight (n=438) |
| --- | --- | --- | --- |
| Session 1 | 48 | 4.23% | 11.0% |
| Session 2 | 71 | 6.26% | 16.2% |
| Session 3 | 95 | 8.37% | 21.7% |

*Cohort-percentage column corrected from 4.0% / 5.9% / 7.9% (calculated against n=1,200) to 4.23% / 6.26% / 8.37% (against n=1,135). The underweight-subgroup-percentage column, calculated against n=438, was correct as published and is unchanged.*

### Summary across all parameters

Table 7 consolidates the corrected outcome data for all five parameters. Improvement rates increase monotonically from session one to session three for every parameter without exception. The largest absolute gain between consecutive sessions occurs between session one and session two across all parameters, consistent with the second session being a critical point for behaviour-change consolidation in this programme, though this observation is descriptive and not tested statistically.

**Table 7.** Consolidated cumulative improvement rates across all five outcome parameters (n=1,135), all percentages recalculated against the analysis cohort.

| Outcome parameter | Session 1 n (%) | Session 2 n (%) | Session 3 n (%) |
| --- | --- | --- | --- |
| BMI improved | 14 (1.23%) | 35 (3.08%) | 44 (3.88%) |
| Waist circumference reduced | 10 (0.88%) | 27 (2.38%) | 39 (3.44%) |
| Hip circumference reduced | 7 (0.62%) | 26 (2.29%) | 34 (3.00%) |
| Waist-to-hip ratio improved | 7 (0.62%) | 25 (2.20%) | 33 (2.91%) |
| Haemoglobin improved | 21 (1.85%) | 73 (6.43%) | 115 (10.13%) |
| Underweight BMI improved | 48 (4.23%) | 71 (6.26%) | 95 (8.37%) |

## DISCUSSION

This re-analysis offers three substantive, and one methodological, contributions to the evidence base for community pharmacist-led health interventions.

The first substantive contribution is that a structured three-session counselling programme delivered by pharmacists was associated with simultaneous, measurable change across five distinct nutritional outcome parameters within three months. Covering both body composition measures and haematological status within a single programme and analysis, this breadth has rarely appeared in the community pharmacy literature and adds descriptive support to the case for expanding what pharmacists are commissioned to do within India’s non-communicable disease prevention system — though, as discussed below, this is associative evidence from a single-arm design, not a demonstration of effect.

The second contribution is the haemoglobin trajectory, the most notable finding in the dataset. An approximately 5.5-fold increase in the proportion of anaemic participants showing measurable haemoglobin improvement, occurring alongside dietary counselling and in the absence of any pharmaceutical product, is consistent with evidence from iron nutrition education programmes in comparable low- and middle-income settings.[16,17] It also sits alongside the broader intervention evidence synthesised by Bhutta et al.,[12] which shows that brief, well-designed nutritional programmes can coincide with biological change even without supplementation, when delivered consistently and adapted to context. This paper cannot establish that the counselling caused the haemoglobin change, given the absence of a control group and any dietary-intake or adherence data; it can only report that the two occurred together.

The third contribution is methodological, and is the central correction in this version of the paper: the original reading of the age-stratified data, in which adolescents aged 11–20 years appeared to be the most responsive subgroup across every parameter, is not supported once the analysis accounts for the fact that this age band comprised 92.4% of the cohort. Rate-adjusted comparison (Table 5), tested formally with Fisher’s exact test, shows that participants aged 21–60 years, though few in number, had statistically significantly higher per-capita improvement rates for BMI, waist circumference, and haemoglobin (p≤0.002 for each). This is the opposite of the original conclusion, and it changes the practical implication of the paper: these data do not support prioritising adolescent-only, school-based delivery on responsiveness grounds. They do support continuing to include adolescents, who make up the large majority of this and likely similar community cohorts, but the case for adolescent-specific targeting needs to rest on other grounds (feasibility of school-based delivery, long-term prevention value, the age structure of the target population) rather than on a claim of superior responsiveness that this dataset does not support.

The pattern in which the largest gain between consecutive sessions falls between session one and session two, consistent across all five parameters, is compatible with the Transtheoretical Model of behaviour change, in which the transition from contemplation to preparation is a stage of high leverage for reinforcement.[19] This is offered as a plausible interpretation rather than a tested one; no formal test of this pattern was conducted. If borne out in future controlled work, it would imply that retention efforts should concentrate on ensuring participants return for session two.

The barriers reported by non-participants — lack of time and lack of interest, each cited by 60% of those who declined — reflect constraints that health information alone is unlikely to address. Integrating counselling into existing school timetables, offering digital reinforcement between sessions, and involving family units rather than individuals in isolation are approaches with supporting evidence.[20] Each is deliverable within infrastructure that Indian community pharmacies and schools already have.

Several limitations should be stated plainly. First, the author was a co-investigator on the primary study; this paper is a re-analysis by a study team member, not an independent evaluation, and should be weighed accordingly — a fact not disclosed in an earlier version of this manuscript and corrected here. Second, the source data record only whether any positive change occurred at each session, not its magnitude, so effect sizes cannot be calculated and no inferential statistics are reported; comparison with randomised trial evidence is correspondingly limited. Third, the absence of a concurrent control group means natural growth trajectories in this predominantly adolescent cohort cannot be separated from any effect of counselling. Fourth, the rate-adjusted age comparison in this paper, while statistically tested and internally consistent, is limited by the unavailability of per-decade group sizes for participants aged 21–60, so the result should be read as evidence that the original adolescent-responsiveness claim is not supported, rather than as proof that any specific older age band responds best. Fifth, no formal test could be applied to the session-to-session improvement trend, because the published data are cumulative counts from the same cohort over time rather than independent samples at each session; testing the true trajectory would require individual-level repeated-measures data that are not available.

Future studies in this population should use a matched control design, record individual-level outcome data with standard deviations to permit inferential analysis, recruit a more age-balanced sample or at minimum report exact denominators for every age band, and extend follow-up to at least six months to establish durability. Biochemical assessment of dietary iron intake would strengthen mechanistic interpretation of the haemoglobin findings.

## CONCLUSION

Three monthly pharmacist-led nutritional counselling sessions were associated with progressive, cumulative improvements in BMI, waist circumference, hip circumference, waist-to-hip ratio, and haemoglobin concentration across a large community-dwelling cohort in India, once outcome percentages are calculated consistently against the analysis cohort (n=1,135). Haemoglobin showed the largest relative change, with an approximately 5.5-fold increase in the proportion of anaemic participants showing measurable improvement, occurring alongside dietary guidance alone. Response was concentrated among adolescents in absolute terms, but this reflects the cohort’s age composition rather than demonstrated differential responsiveness; rate-adjusted analysis suggests the reverse may be true, though the available data cannot confirm this. These findings offer descriptive, hypothesis-generating support for continued evaluation of pharmacist-led counselling in community-based noncommunicable disease prevention in India. Future research should prioritise randomised or controlled designs, individual-level outcome measurement, age-balanced recruitment with complete reporting of subgroup denominators, and extended follow-up.

## Data Availability

All data analysed in this study are available in the primary publication: Suresh Kumar JN et al. A prospective interventional study on prevalence of obesity and anaemia in community settings. IOSR Journal of Pharmacy. 2023;13(5):15-34. Available at: http://www.iosrphr.org/papers/vol13-issue5/C1305011534.pdf

## DECLARATIONS

Ethics approval and consent to participate: The primary study received ethics approval from the Institutional Ethics Committee of Narasaraopeta Institute of Pharmaceutical Sciences (IEC-NIPS/PPP/2021-22/003). This re-analysis draws only on aggregate data already collected under that approval and does not involve new access to individual participant records. Separate ethics approval was therefore not required.

### Consent for publication

Not applicable, as no individual participant data are reported in this paper.

### Availability of data and materials

All source data used in this analysis are available in the primary publication: Suresh Kumar JN, Bhavya Sai S, Naga Amulya D, Ritheesha D, Priyanka KSL, Chaitanya Kumar S. A prospective interventional study on prevalence of obesity and anaemia in community settings. IOSR Journal of Pharmacy. 2023;13(5):15-34. The extracted counts underlying every table in this paper, together with the Wilson confidence interval and Fisher’s exact test calculations for the age-band comparison, are provided in the accompanying Supplementary Data spreadsheet.

### Funding: None

The primary study also received no external funding.

### Authors’ contributions

RD was a co-investigator on the primary study team that generated the source dataset (Suresh Kumar JN, Bhavya Sai S, Naga Amulya D, Ritheesha D, Priyanka KSL, Chaitanya Kumar S), and independently carried out the conceptualisation, denominator correction, rate-adjusted analytical framework, interpretation, literature synthesis, and manuscript drafting and revision for this re-analysis.

## Acknowledgements

The author thanks the research team at Narasaraopeta Institute of Pharmaceutical Sciences for generating the primary dataset that made this analysis possible, and the 1,200 community participants whose contributions underpin the findings reported here.

